# Acceptability of Doxy-PEP Among Men Who Have Sex With Men in New York City: Findings From the 2023 National HIV Behavioral Surveillance Survey

**DOI:** 10.64898/2026.05.28.26354355

**Authors:** Étienne Meunier, Alexis Rivera, Kristina Rodriguez, Pablo A. Martinez, Zoe Edelstein

## Abstract

In the United States, gay, bisexual, and other men who have sex with men (MSM) experience a disproportionate burden of sexually transmitted infections (STIs), with notable racial/ethnic disparities. Doxycycline post-exposure prophylaxis (doxy-PEP) has emerged as a promising strategy to prevent bacterial STIs. This study analyzed 2023 National HIV Behavioral Surveillance data to examine doxy-PEP awareness, use, and intent to use among MSM in New York City (NYC), in a predominantly Hispanic/Latino sample. Among 134 participants, awareness and prior use were low (38.8% and 9.0%, respectively), but intent to use was high (75.4%). In Poisson regression models, intent was higher among participants reporting non-injection drug use and 2–10 partners in the past 12 months, while marginally lower among those with lower incomes and recent migrants. Findings suggest doxy-PEP is acceptable for MSM in NYC, but addressing barriers among low-income groups and recent migrants is critical to reducing disparities.

## Introduction

In the United States (US), gay, bisexual, and other men who have sex with men (MSM) are disproportionately impacted by sexually transmitted infections (STIs). The Centers for Disease Control and Prevention (CDC) estimated that, in 2023, MSM accounted for one-third of all syphilis cases and half of gonorrhea cases [1]. In New York City (NYC) in 2024, men represented more than 80% of gonorrhea and syphilis diagnoses, with syphilis rates among Black and Hispanic/Latino men approximately three times higher than among White men [2].

Doxycycline post-exposure prophylaxis (doxy-PEP) has emerged as a promising strategy to prevent STIs. Clinical trials among MSM have shown that a 200 mg dose of doxycycline within 72 hours of sex reduces incident bacterial STIs, with greater efficacy for chlamydia and syphilis than gonorrhea [3]. In 2024, the CDC issued guidance recommending that providers discuss doxy-PEP with MSM who had a bacterial STI in the past year [4].

Before the CDC guidance, studies among MSM in various U.S. settings found doxy-PEP awareness was low, yet interest was high [5–9]. Our study offers early evidence on doxy-PEP acceptability in NYC among a sample predominantly composed of Hispanic/Latino MSM, a population with a high burden of STIs and limited prior-research representation.

## Methods

Data were collected as part of the 2023 MSM cycle of the CDC’s National HIV Behavioral Surveillance (NHBS) study, a cyclical cross-sectional survey conducted in 19 U.S. cities (the full protocol is available online [10]). This analysis draws on data from the NYC site. Participants were recruited using venue-based, time-space sampling. Eligibility criteria included aged 18 or older, assigned male at birth, identifying as man or nonbinary, having ever had oral or anal sex with another man, fluency in English or Spanish, and residence in the NYC metropolitan area. Participants completed an anonymous interviewer-administered questionnaire and were offered HIV testing. The NYC Health Department’s Institutional Review Board approved procedures, and all participants provided informed consent. We offered gift-card incentives of $50 for completing the survey and $50 for HIV testing.

The main outcomes were doxy-PEP awareness, lifetime use, and intent to use. Interviewers read a brief definition of doxy-PEP and asked if participants had ever heard of it and, if so, had ever used it. Then, all participants answered on a 5-point scale “In the future, how likely would you be to use doxy-PEP if it was available to you?” We grouped responses into intent to use (“definitely yes” or “probably yes”) versus no intent (“might or might not,” “probably not,” or “definitely not”). Other variables included demographics, sexual behaviors, substance use, health care access, and sexual-health service use.

We used cross-tabulations to describe the outcome variables. To identify factors associated with awareness and intent, we used log-linked Poisson regression models with robust standard errors. Variables with *p*-value < 0.1 in unadjusted models were included in adjusted models. To avoid multicollinearity, we excluded correlated variables capturing overlapping constructs (e.g., marijuana and non-injection drug use). We refined models using backward stepwise selection to improve fit and prevent overfitting due to sample size. Analyses were conducted in IBM SPSS Statistics 30.

## Results

Table 1 presents characteristics for the 134 MSM enrolled. Most identified as Hispanic/Latino (61.9%) and 63.4% were born outside the US or Puerto Rico (91.8% of them came from Latin America). Over two-thirds (67.9%) were aged 30 or older, while only half (50.0%) reported at least some college-level education. More than a third (36.4%) lived at or below the 2023 U.S. Federal Poverty Level (FPL) [11], and nearly one in five (18.7%) experienced homelessness in the past 12 months. Most identified as cisgender and as gay, with only five (3.7%) identifying as nonbinary and 11 (8.2%) as bisexual.

**Table 1.** Doxy PEP awareness, use, and intent to use by characteristics of interest among men who have sex with men, National HIV Behavioral Surveillance Study, New York City, 2023 (n = 134).

|  | Total |  | Awareness |  | Use |  | Intent to use |  |
| --- | --- | --- | --- | --- | --- | --- | --- | --- |
|  | *n* | (%) | *n* | (%) | *n* | (%) | *n* | (%) |
|  | 134 | (100.0) | 52 | (38.8) | 12 | (9.0) | 101 | (75.4) |
| **Demographics** |  |  |  |  |  |  |  |  |
| 30 years or older | 91 | (67.9) | 42 | (80.8) | 9 | (75.0) | 74 | (73.3) |
| Race/ethnicity |  |  |  |  |  |  |  |  |
| Hispanic/Latino | 83 | (61.9) | 31 | (59.6) | 8 | (66.7) | 59 | (58.4) |
| Black | 16 | (11.9) | 5 | (9.6) | 0 | (0.0) | 13 | (12.9) |
| Other | 35 | (26.1) | 16 | (30.8) | 4 | (33.3) | 29 | (28.7) |
| Identifies as man | 129 | (96.3) | 51 | (98.1) | 12 | (100.0) | 97 | (96.0) |
| Identifies as gay | 123 | (91.8) | 48 | (92.3) | 12 | (100.0) | 93 | (92.1) |
| Time in the US |  |  |  |  |  |  |  |  |
| Born in the US | 49 | (36.6) | 18 | (34.6) | 4 | (33.3) | 38 | (37.6) |
| Moved to US before 2022 | 48 | (35.8) | 28 | (53.8) | 8 | (66.7) | 43 | (42.6) |
| Moved to US in 2022 or more recently | 37 | (27.6) | 6 | (11.5) | 0 | (0.0) | 20 | (19.8) |
| Lives at or below 2023 Federal Poverty Level | 48 | (36.4) | 13 | (25.0) | 4 | (33.3) | 28 | (28.3) |
| Homelessness<sup>a</sup> | 25 | (18.7) | 5 | (9.6) | 0 | (0.0) | 15 | (14.9) |
| Some college-level education or more | 67 | (50.0) | 32 | (61.5) | 9 | (75.0) | 54 | (53.5) |
| **Sexual behaviors** |  |  |  |  |  |  |  |  |
| Number of sex partners<sup>a</sup> |  |  |  |  |  |  |  |  |
| 1 | 18 | (13.4) | 2 | (3.8) | 0 | (0.0) | 10 | (9.9) |
| 2–10 | 65 | (48.5) | 28 | (53.8) | 8 | (66.7) | 55 | (54.5) |
| ≥10 | 51 | (38.1) | 22 | (42.3) | 4 | (33.3) | 36 | (35.6) |
| Group sex venue attendance<sup>a</sup> | 72 | (53.7) | 31 | (59.6) | 7 | (58.3) | 53 | (52.5) |
| **Substance use behaviors** |  |  |  |  |  |  |  |  |
| Non-injection drug use<sup>a,b</sup> | 37 | (27.6) | 17 | (32.7) | 5 | (41.7) | 32 | (31.7) |
| Marijuana use<sup>a</sup> | 64 | (47.8) | 26 | (50.0) | 7 | (58.3) | 53 | (52.5) |
| Binge alcohol use<sup>c</sup> | 38 | (28.4) | 12 | (23.1) | 3 | (25.0) | 29 | (28.7) |
| **Health care access and use of sexual health services** |  |  |  |  |  |  |  |  |
| Currently insured | 113 | (85.0) | 46 | (88.5) | 10 | (83.3) | 89 | (88.1) |
| Had a health care visit<sup>a</sup> | 120 | (89.6) | 46 | (88.5) | 9 | (75.0) | 93 | (92.1) |
| Received STI testing<sup>a</sup> | 98 | (73.1) | 38 | (73.1) | 7 | (58.3) | 79 | (78.2) |
| Had a bacterial STI diagnosis<sup>a</sup> | 48 | (35.8) | 18 | (34.6) | 5 | (41.7) | 39 | (38.6) |
| Aware of HIV PrEP | 121 | (90.3) | 49 | (94.2) | 12 | (100.0) | 94 | (93.1) |
| Ever used HIV PrEP | 78 | (58.2) | 34 | (65.4) | 9 | (75.0) | 64 | (63.4) |
| Self-reported having HIV | 37 | (27.6) | 12 | (23.1) | 2 | (16.7) | 27 | (26.7) |
| Received an HIV test<sup>a,d</sup> | 83 | (85.6) | 36 | (90.0) | 10 | (100.0) | 64 | (86.5) |
| Used HIV PEP<sup>a,d</sup> | 13 | (13.4) | 4 | (10.0) | 0 | (0.0) | 9 | (12.2) |
| Used HIV PrEP<sup>a,d</sup> | 66 | (68.0) | 31 | (77.5) | 8 | (80.0) | 54 | (73.0) |

Abbreviations: PEP: post-exposure prophylaxis; STI: sexually transmitted infection; PrEP: pre-exposure prophylaxis.
<sup>a</sup> In the past 12 months
<sup>b</sup> Used any drugs other than marijuana that were not prescribed and not injected
<sup>c</sup> In the past 30 days, 5 or more standard drinks within about 2 hours
<sup>d</sup> Among those who did not self-report having HIV (n = 97)

Approximately two in five (38.8%) reported prior awareness of doxy-PEP, while few (9.0%) had ever used it. Intent to use was high: three-quarters (75.4%) indicated they would probably or definitely use doxy-PEP in the future.

Table 2 presents unadjusted and adjusted analyses. We could not examine associations with prior use as only 12 participants reported it. In unadjusted analyses, those who were aware of doxy-PEP were more likely to be aged 30 or older and to have multiple sex partners of any gender in the past 12 months, while they were less likely to have moved to the US in 2022 or after and to live at or below the FPL. After adjustment, awareness was significantly lower among participants who moved to the US in 2022 or later (aPR = 0.40; 95% CI: 0.18– 0.86) and lower with marginal significance (*p* < .1) among those reporting only one partner in the past 12 months.

**Table 2.**
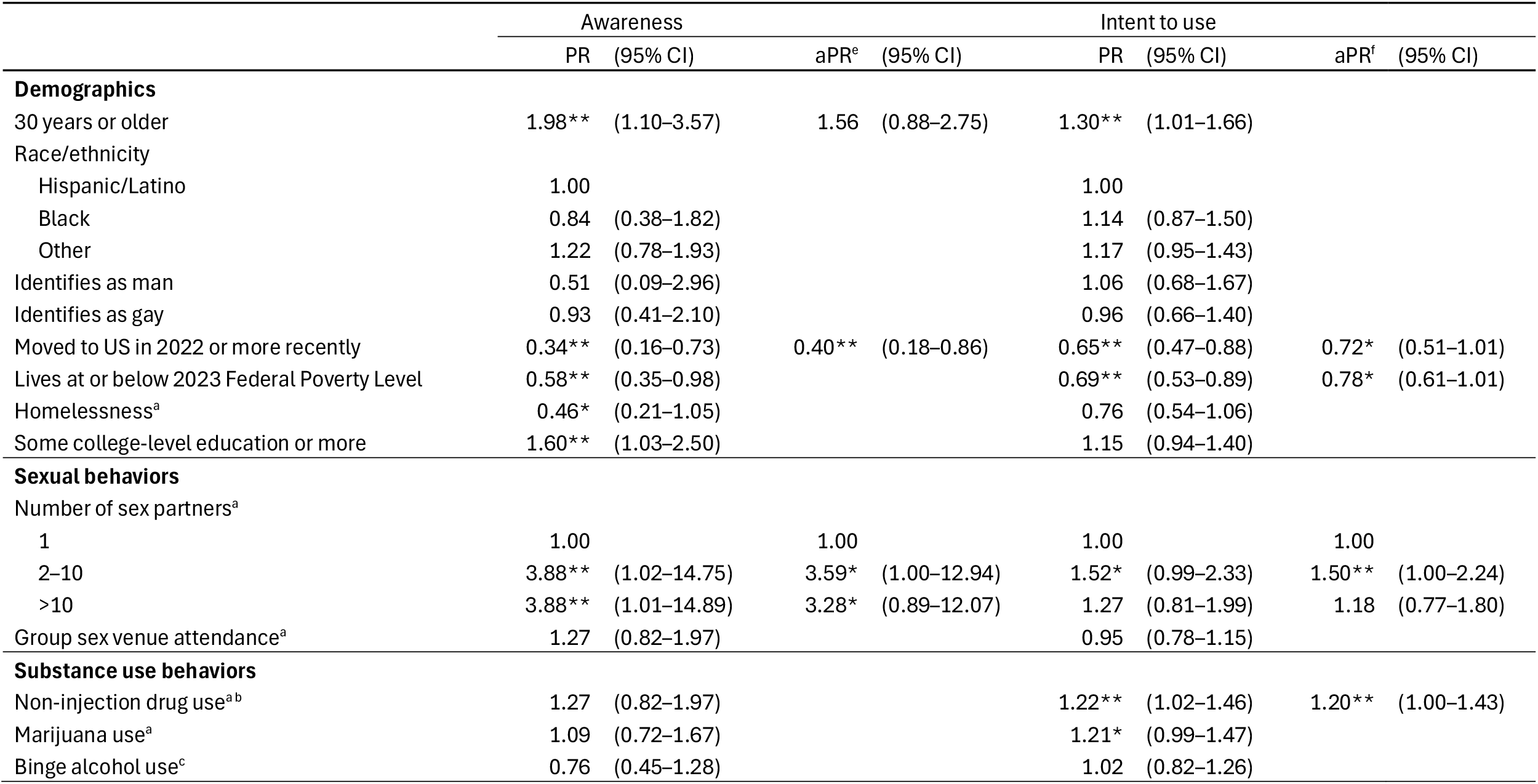
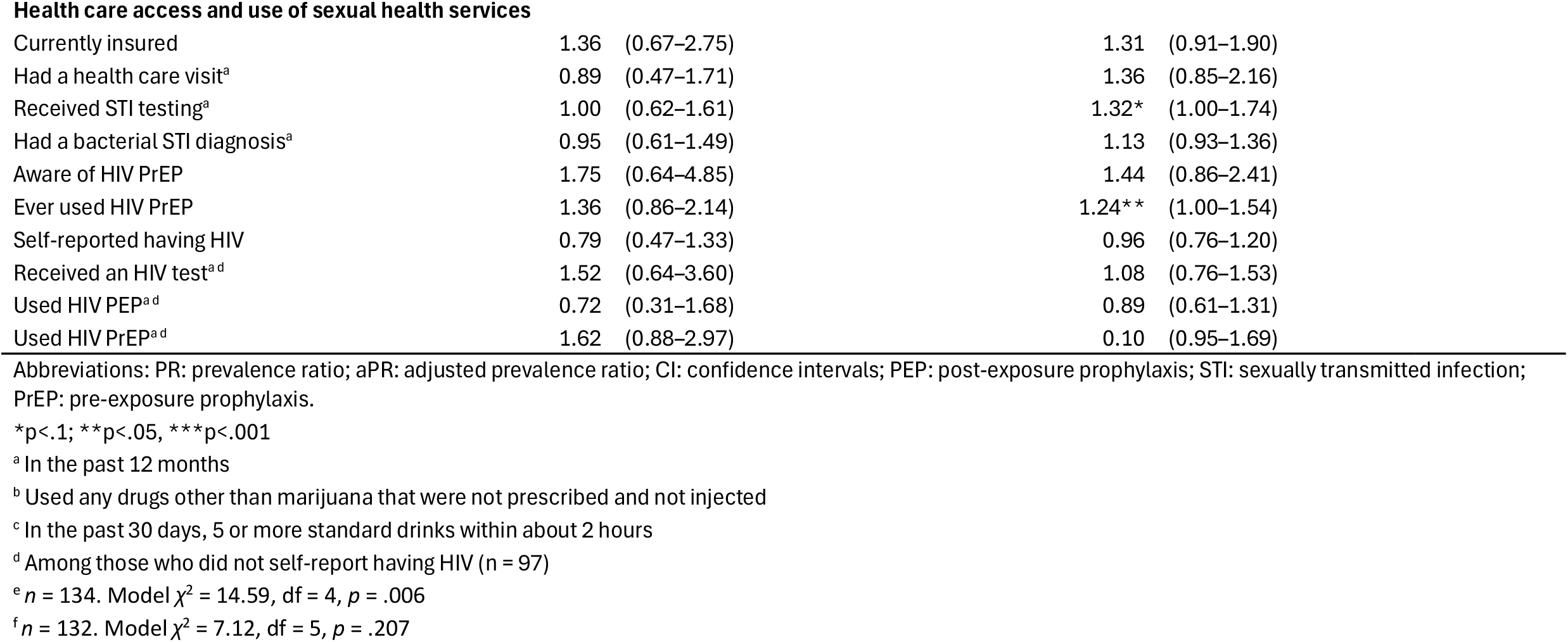
Associations of Doxy PEP interest among NHBS MSM 2023 participants.

In unadjusted analyses, those who reported intent to use doxy-PEP were more likely to be aged 30 or older, have been in the US before 2022, live above the FPL, report 2–10 sex partners, use non-injection drugs, and have ever used PrEP to prevent HIV. After adjustment, participants reporting non-injection drug use in the past 12 months were more likely to intend to use doxy-PEP (aPR = 1.20; 95% CI: 1.00–1.43). Intent was also higher among participants reporting 2–10 sex partners in the prior 12 months compared to those who reported only one (aPR = 1.50; 95% CI: 1.00–2.24), though it did not differ significantly with those reporting more than 10 partners. Participants who arrived in the US in 2022 or later (aPR = 0.72; 95% CI: 0.51–1.01) and those living at or below the FPL (aPR = 0.78; 95% CI: 0.61–1.01) were marginally less likely to intend to use doxy-PEP.

## Discussion

In this survey, we found low uptake of doxy-PEP among MSM in NYC in 2023, as only 38.8% had heard of it and 9.0% had ever used it. These figures align with another 2023 survey of patients at NYC Sexual Health Clinics, where 28.9% reported awareness and 11.1% prior use [12]. The low awareness and use could be expected as the CDC and local health departments released doxy-PEP guidance only toward the end of 2023 [13]. In contrast, the 2023 San Francisco NHBS-MSM sample reported substantially higher awareness (66.6%) and prior use (19.3%) [9], likely reflecting that city’s earlier adoption of doxy-PEP recommendations in 2022. Awareness and use have likely risen with new provider guidance and expanded promotion.

Despite limited awareness, 75.4% expressed intent to use doxy-PEP in the future, consistent with similar studies [5]. Given that most participants were Hispanic/Latino MSM, a group disproportionately affected by STIs in NYC [2], this high acceptability highlights doxy-PEP’s potential to reduce disparities. However, awareness was lower among participants who arrived in the US in 2022 or later, and intent was lower among them and those living below the FPL. While we did not ask the reasons for intent, results may reflect challenges navigating the healthcare system for recent migrants [14]. Prior studies identified anticipated costs as a common barrier to doxy-PEP use [12,15,16], which may help explain reduced interest among low-income groups. Some may be unaware doxycycline is a relatively inexpensive generic medication or worry about costs related to medical visits. Addressing these concerns is especially important because STI diagnosis rates in NYC are highest in high-poverty areas [2]. To reduce such disparities, it will be essential to ensure doxy-PEP is affordable or free and that priority populations are aware of it.

Participants reporting non-injection drug use in the past 12 months were more likely to intend to use doxy-PEP, consistent with other studies showing higher interest among people who use substances [17,18]. Some may engage in drug use during sex, which research has linked to increased STI transmission [19]. As intoxication can hinder other prevention strategies like condom use, MSM who use substances may view doxy-PEP as a complementary prevention tool, and it should be integrated into sexual-health services for this population.

Prior studies showed mixed results regarding the relationship between number of sex partners and doxy-PEP acceptability [17,20]. In our sample, intent was only higher among those with 2–10 partners, not those reporting more than 10. These findings should be interpreted cautiously due to the small convenience sample but raise interesting questions about doxy-PEP acceptability among highly sexually active individuals. Antibiotic resistance and side effects are commonly reported barriers to doxy-PEP use [12,15,16], and there could be concerns about frequent dosing among people with high number of sex partners. Future research should not only explore intent to use but also how often and under what circumstances people would use doxy-PEP.

The small, venue-based sample limits the generalizability of these findings. Interviewer-administered questionnaires can introduce social-desirability bias, leading participants to report favorable attitudes toward STI prevention strategies like doxy-PEP. Behavioral data were self-reported and are subject to recall error.

Despite these limitations, our survey found patterns similar to other studies and populations. The results are encouraging for reducing STI disparities among Hispanic/Latino MSM in NYC, while pointing to challenges in doxy-PEP uptake among recent migrants and low-income groups that public-health workers and clinical providers will need to overcome.

## Declarations

### Ethics approval

The study presented in this article was approved by the Institutional Review Board at the New York City Department of Health and Mental Hygiene (protocol #04-006; PI: Rivera).

### Consent

All individuals provided informed consent prior to participating in the study presented in this article.

### Data availability

Data from this study are not publicly available. Please contact the authors with specific inquiries.

### Authors’ contribution

AR, KR, and PM participated in study design, implementation, and data collection. EM led the analysis and wrote the initial version of the manuscript. All authors provided feedback on the analysis. All authors reviewed drafts and approved the final version of the manuscript.

## Acknowledgements

The authors would like to acknowledge NHBS-MSM2023 data collection staff and study participants.

